# Second multistate outbreak of tuberculosis caused by a bone allograft product

**DOI:** 10.64898/2026.04.29.26351868

**Authors:** Kimberly R. Schildknecht, Paula M. Williams, Noah G. Schwartz, Maryam B. Haddad, Rebekah J. Stewart, Pallavi Annambhotla, Sridhar V. Basavaraju, Scott A. Nabity, Chris E. Keh, Helene M. Calvet, Matthew M. Zahn, Romina Beltrán, Ashley Cortez, Andrea Lomeli, Jeffrey M. Percak, Lisa L. Goozé, Myron Coloma, Tambi Shaw, Peter J. Davidson, Shona R. Smith, Robert P. Dickson, Daniel R. Kaul, Annett R. Gonzalez, Gretchen Rodriguez, Amanda Decimo, Audilis Sanchez, Lisa Y. Armitige, Jessica Stapleton, Michael Lacassagne, Chris Brown, Crystal Zheng, Juzar Ali, Amy W. Wolfe, Laura R. Young, Kiley Ariail, Heidi Behm, Hannah T. Jordan, Magdalene Spencer, Diana M. Nilsen, RuthAnn Goradia, Brenda Montoya Denison, Marcos Burgos, Juliet M. Leonard, Erick Cortes, Tyler C. Thacker, Kimberly A. Lehman, Julian A. Villalba, Julu Bhatnagar, Sarah Reagan-Steiner, Sandy P. Althomsons, Neela D. Goswami, Lauren S. Cowan, Angela M. Starks, Jonathan M. Wortham, Philip A. LoBue, the Bone Allograft Tuberculosis 2023 Investigators

**Affiliations:** Division of Tuberculosis Elimination, National Center for HIV, Viral Hepatitis, STD, and TB Prevention, Centers for Disease Control and Prevention, Atlanta, Georgia, USA; Epidemic Intelligence Service, Centers for Disease Control and Prevention, Atlanta, Georgia, USA; Division of Healthcare Quality Promotion, National Center for Emerging and Zoonotic Infectious Diseases, Centers for Disease Control and Prevention, Atlanta, Georgia, USA; California Department of Public Health, Richmond, California, USA; Orange County Health Care Agency, Santa Ana, California, USA; County of San Diego Health and Human Services Agency, San Diego, California, USA; San Mateo County Health, San Mateo, California, USA; Michigan Department of Health and Human Services, Lansing, Michigan, USA; University of Michigan, Ann Arbor, Michigan, USA; City of El Paso Department of Public Health, El Paso, Texas, USA; Texas Department of State Health Services, Austin, Texas, USA; University of Texas at Tyler Health Science Center, Tyler, Texas, USA; Louisiana Department of Health, Baton Rouge, Louisiana, USA; Tulane University School of Medicine, New Orleans, Louisiana, USA; Louisiana State University Health Sciences Center, New Orleans, Louisiana, USA; Virginia Department of Health, Richmond, Virginia, USA; Oregon Public Health Division, Portland, Oregon, USA; New York City Department of Health and Mental Hygiene, New York City, New York, USA; New Mexico Department of Health, Albuquerque, New Mexico, USA; New Jersey Department of Health, Trenton, New Jersey, USA; National Veterinary Services Laboratories, U.S. Department of Agriculture, Ames, Iowa, USA; Division of High Consequence Pathogens and Pathology, National Center for Emerging and Zoonotic Infectious Diseases, Centers for Disease Control and Prevention, Atlanta, Georgia, USA

## Abstract

Tuberculosis screening is not mandatory for prospective tissue donors. In 2021 and 2023, two different bone allograft products caused nationwide tuberculosis outbreaks. We assessed the morbidity and mortality of the second outbreak and reviewed donor and tissue screening to identify deficiencies. Thirty-six people residing in nine states received the product during spinal and dental procedures. Twenty-seven recipients had tuberculosis infection, 11 had microbiologic or imaging evidence of tuberculosis disease, and two died from tuberculosis within 12 months of outbreak detection. Another recipient died from tuberculosis nearly 3 years after product implantation. The bone donor died of pneumonia and septic shock. Polymerase chain reaction testing of the product before and after distribution did not detect *Mycobacterium tuberculosis*. Mycobacterial culture was not performed until after outbreak detection, when *M. tuberculosis* was isolated from 2 of 6 unused product units. This outbreak demonstrates persistent gaps in tissue transplant safety. Appropriate selection of donors and mycobacterial culture of donated tissues could reduce but not eliminate the risk of *M. tuberculosis* transmission. Therefore, it is important that clinicians monitor tissue recipients and promptly report adverse events to tissue establishments and health authorities.

## 1 Introduction

In 2021, a bone allograft product from a deceased donor with unrecognized tuberculosis disease was implanted into 113 people, causing a multistate outbreak that led to multiple deaths.^1^ In 2023, a bone allograft product from a different deceased donor (i.e., with a unique lot number) caused a second nationwide tuberculosis outbreak, again leading to deaths in recipients.^2^ The initial public health response to the second outbreak and clinical features of several cases have been described.^2–4^ Here, we comprehensively describe morbidity and mortality among all product recipients linked to the second outbreak and identify deficiencies in donor selection and tissue testing.

## 2 Materials and methods

### 2.1 Identification of outbreak cohort

During February 27–June 20, 2023, 50 units from the affected product lot were shipped to 8 hospitals and 5 dental facilities in 7 states (Supplementary Figure 1). The Centers for Disease Control and Prevention (CDC) worked with the product manufacturer (i.e., the tissue establishment), health departments, and healthcare facilities to account for all units by July 14, 2023, 1 week after the first tuberculosis case notification.^2^ During February 28–June 22, 49 of the 50 units had been implanted into 36 recipients residing in 9 states (some received ≥1 unit); the 50^th^ unit was unused and successfully sequestered (Figure 1). An additional 53 units not yet distributed by the manufacturer were also sequestered. Immediately after outbreak detection in July 2023, CDC recommended that all recipients begin empiric multidrug treatment for tuberculosis disease.^2^

**Figure 1.**
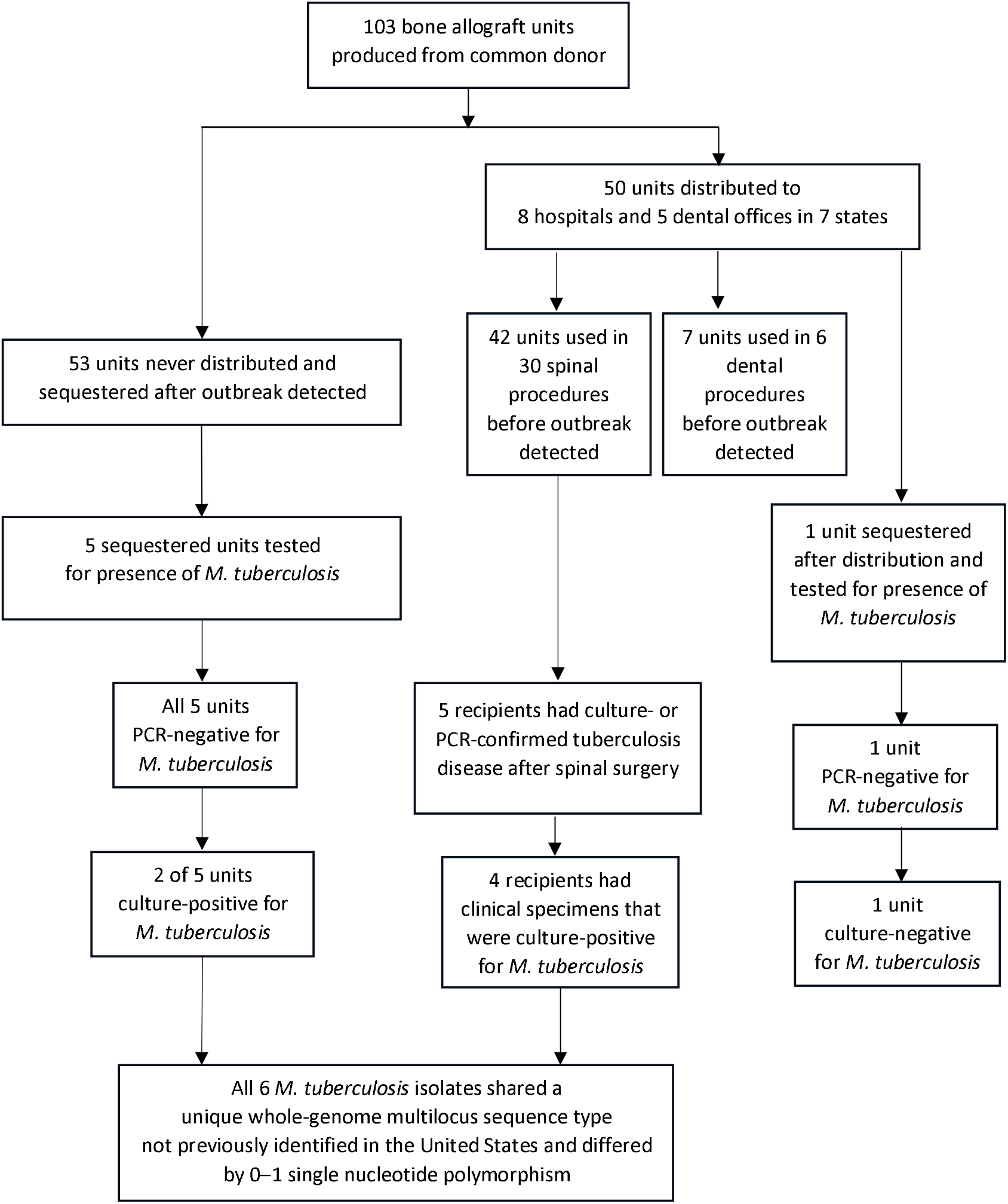
Flowchart showing testing of bone allograft recipients and unused units for *Mycobacterium tuberculosis*. In total, 103 bone allograft units were produced; 54 units were sequestered before use, and 49 units were implanted into 36 recipients. Six unused units were tested, with all 6 PCR-negative but 2 culture-positive. During the 12 months after outbreak detection, 5 recipients developed culture- or PCR-confirmed tuberculosis, of whom 4 had culture-positive specimens. All 6 *M. tuberculosis* isolates from unused units and recipients shared a unique whole-genome multilocus sequence type not previously identified in the United States and differed by 0–1 single nucleotide polymorphism. After the end of cohort follow-up, one additional recipient developed culture-confirmed tuberculosis with an isolate sharing the same whole-genome multilocus sequence type. M. tuberculosis, Mycobacterium tuberculosis; PCR, polymerase chain reaction.

### 2.2 Prospective cohort study to assess morbidity and mortality

During July 2023–July 2024, we conducted a prospective cohort study including all 36 product recipients to determine the frequency of tuberculosis infection and disease, hospitalization, repeat surgery, death, and treatment completion. CDC investigators distributed standardized case report forms at outbreak detection and 12 months later. Investigators from state and local health departments extracted demographic, clinical, laboratory, and imaging data from medical records and reported whether there was laboratory or imaging evidence of tuberculosis infection or disease. If any data elements were missing from case report forms, equivalent data were obtained from tuberculosis surveillance forms routinely submitted by health departments to CDC.^5^

Recipients were classified as having microbiologically confirmed tuberculosis disease if *Mycobacterium tuberculosis* complex was detected in a clinical specimen by mycobacterial culture or nucleic acid amplification testing (NAAT), including polymerase chain reaction (PCR). Recipients with positive mycobacterial testing or imaging consistent with tuberculosis disease were classified as having evidence of disease at the following sites: spine or paraspinal soft tissues, dental procedure site, lungs, central nervous system, or other sites. For recipients who died, state health departments reported whether tuberculosis was an underlying or contributing cause of death. CDC’s Infectious Diseases Pathology Branch (Atlanta, GA) performed postmortem pathologic examination of formalin-fixed, paraffin-embedded specimens from spine, brain, and visceral tissue from one recipient, which included immunohistochemical staining, PCR testing (targeting the IS*6110* insertion sequence and *16S rRNA* gene), and Sanger sequencing of positive amplicons in DNA extracts. Deidentified data were stored in a secure REDCap database.^6^ Descriptive statistics were calculated using SAS version 9.4 (SAS Institute Inc).

### 2.3 Investigation of donor and tissue screening

CDC and the U.S. Food and Drug Administration (FDA) worked with the tissue recovery firm and product manufacturer to review the donor’s medical history, screening and testing records, and tissue recovery and processing procedures. Unused units from the product lot were evaluated at the U.S. Department of Agriculture National Veterinary Services Laboratories (Ames, IA) for the presence of *M. tuberculosis* complex using a real-time PCR assay that amplifies the IS*1081* insertion element, and by mycobacterial culture in liquid and solid media. To determine the genetic relatedness of *M. tuberculosis* isolates from unused product and product recipients, the National Tuberculosis Molecular Surveillance Center (Lansing, MI) performed whole-genome sequencing, and CDC performed whole-genome multilocus sequence typing and phylogenetic analysis. Detailed laboratory methods are available in Supplementary Methods.

### 2.4 Investigation oversight

This investigation was reviewed by CDC, deemed not to constitute research, and conducted consistent with applicable federal law and CDC policy (45 C.F.R. part 46, 21 C.F.R. part 56; 42 U.S.C. Sect. 241(d); 5 U.S.C. Sect. 552a; 44 U.S.C. Sect. 3501 et seq).

## 3 Results

### 3.1 Baseline characteristics of product recipients

Thirty of the 36 recipients had undergone spinal surgeries (Table 1) with 1–4 units (10 cc per unit) of the bone allograft product. The other 6 recipients had undergone dental procedures with 1–2 units (1 cc per unit) of the bone allograft product. The most common site of spinal surgeries was lumbosacral (22 of 30 [73%]), and all 6 dental procedures were performed on the maxilla.

**Table 1.** Baseline characteristics of 36 patients who received a bone allograft product containing *Mycobacterium tuberculosis*, total and by procedure type.

| Characteristics | No./Total (%)<sup>a</sup> |  |  |
| --- | --- | --- | --- |
|  | All recipients (N = 36) | Spinal recipients (n = 30) | Dental recipients (n = 6) |
| Age, median (range), y | 59 (33–82) | 60 (36–82) | 45 (33–80) |
| Female sex | 21/36 (58%) | 17/30 (57%) | 4/6 (67%) |
| Born in the United States<sup>b</sup> | 32/36 (89%) | 27/30 (90%) | 5/6 (83%) |
| Obesity (body mass index $\geq 30$ kg/m<sup>2</sup>) | 16/36 (44%) | 16/30 (53%) | 0 |
| Medical risk factors for tuberculosis |  |  |  |
| Diabetes mellitus | 8/36 (22%) | 8/30 (27%) | 0 |
| Immunocompromising condition or medication<sup>c</sup> | 7/36 (19%) | 7/30 (23%) | 0 |
| Liver disease | 3/36 (8%) | 3/30 (10%) | 0 |
| End-stage renal disease | 2/36 (6%) | 2/30 (7%) | 0 |
| Site of bone allograft implantation |  |  |  |
| Cervical spine | 6/36 (17%) | 6/30 (20%) | NA |
| Thoracic spine | 1/36 (3%) | 1/30 (3%) | NA |
| Thoracic and lumbosacral spine | 1/36 (3%) | 1/30 (3%) | NA |
| Lumbosacral spine | 22/36 (61%) | 22/30 (73%) | NA |
| Maxilla | 6/36 (17%) | NA | 6/6 (100%) |

NA, not applicable.
<sup>a</sup> Due to rounding, column percentages for site of bone allograft implantation do not sum to 100%.
<sup>b</sup> Born in the 50 U.S. states, the District of Columbia, or Puerto Rico.
<sup>c</sup> One recipient had a history of kidney transplantation, 3 had rheumatoid arthritis, 2 were receiving chronic corticosteroids for unknown indications, and 1 was receiving mepolizumab for severe asthma.

Product recipients were primarily female and born in the United States. The spinal recipients had an older median age of 60 years (range, 36–82 years) compared with dental recipients, whose median age was 45.5 years (range, 33–80 years).

### 3.2 Tuberculosis morbidity, mortality, and treatment outcomes

Among the 35 recipients with post-procedure test results for *M. tuberculosis* infection reported, 27 (77%) had positive interferon-gamma release assay results, including 24 spinal recipients and 3 dental recipients (Table 2). Eight of the 27 had tested negative before the bone allograft implantation, and the others had no known previous tests for *M. tuberculosis* infection.

**Table 2.**
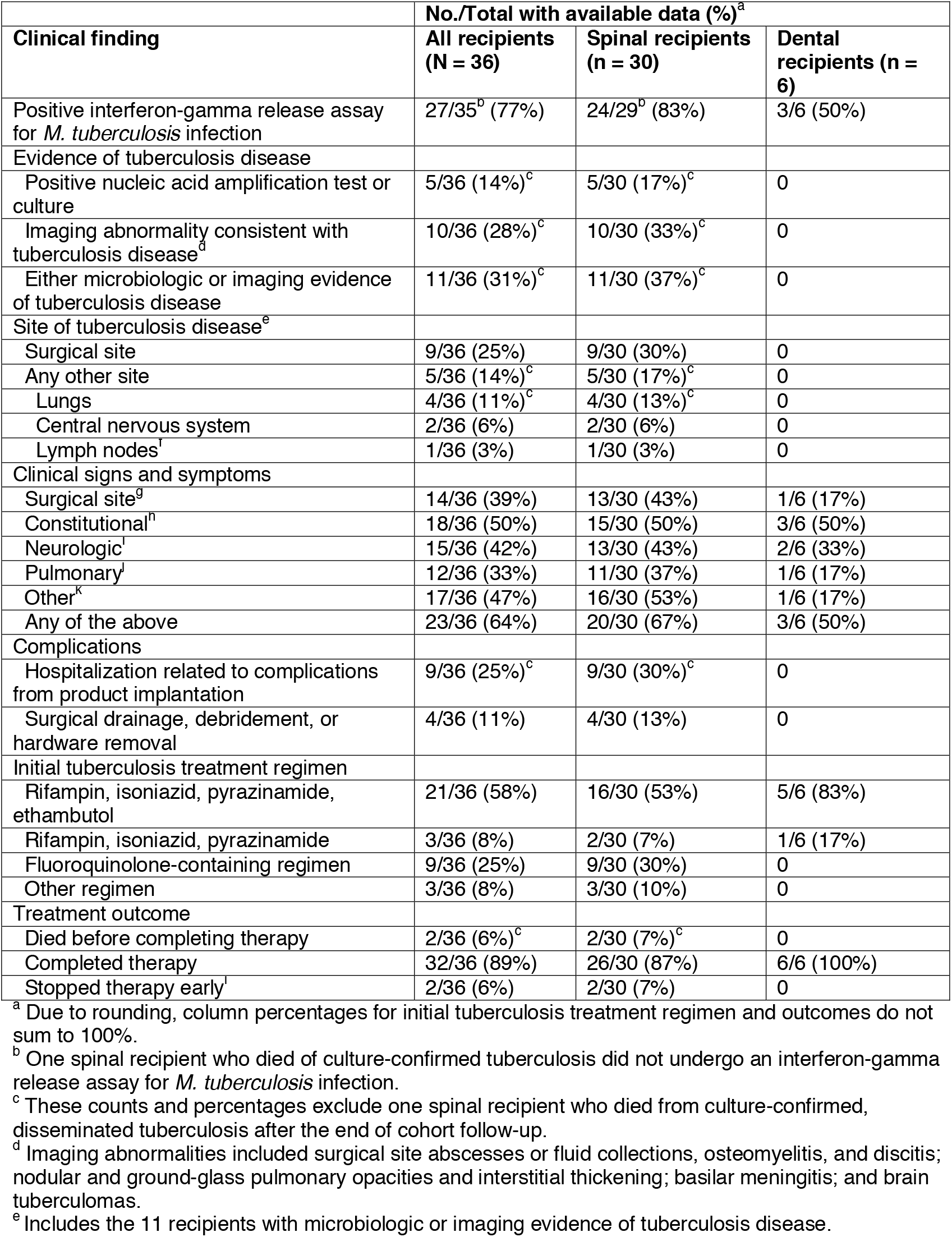
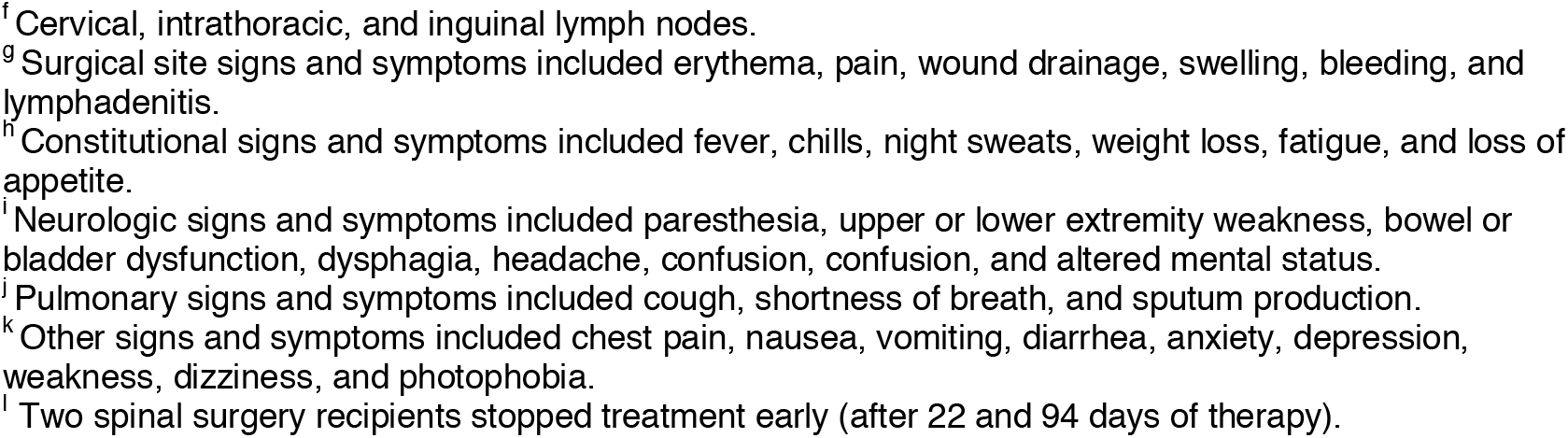
Clinical findings in 36 patients who received a bone allograft product containing *Mycobacterium tuberculosis*, total and by procedure type.

| Clinical finding | No./Total with available data (%)<sup>a</sup> |  |  |
| --- | --- | --- | --- |
|  | All recipients (N = 36) | Spinal recipients (n = 30) | Dental recipients (n = 6) |
| Positive interferon-gamma release assay for *M. tuberculosis* infection | 27/35<sup>b</sup> (77%) | 24/29<sup>b</sup> (83%) | 3/6 (50%) |
| Evidence of tuberculosis disease |  |  |  |
| Positive nucleic acid amplification test or culture | 5/36 (14%)<sup>c</sup> | 5/30 (17%)<sup>c</sup> | 0 |
| Imaging abnormality consistent with tuberculosis disease<sup>d</sup> | 10/36 (28%)<sup>c</sup> | 10/30 (33%)<sup>c</sup> | 0 |
| Either microbiologic or imaging evidence of tuberculosis disease | 11/36 (31%)<sup>c</sup> | 11/30 (37%)<sup>c</sup> | 0 |
| Site of tuberculosis disease<sup>e</sup> |  |  |  |
| Surgical site | 9/36 (25%) | 9/30 (30%) | 0 |
| Any other site | 5/36 (14%)<sup>c</sup> | 5/30 (17%)<sup>c</sup> | 0 |
| Lungs | 4/36 (11%)<sup>c</sup> | 4/30 (13%)<sup>c</sup> | 0 |
| Central nervous system | 2/36 (6%) | 2/30 (6%) | 0 |
| Lymph nodes<sup>f</sup> | 1/36 (3%) | 1/30 (3%) | 0 |
| Clinical signs and symptoms |  |  |  |
| Surgical site<sup>g</sup> | 14/36 (39%) | 13/30 (43%) | 1/6 (17%) |
| Constitutional<sup>h</sup> | 18/36 (50%) | 15/30 (50%) | 3/6 (50%) |
| Neurologic<sup>i</sup> | 15/36 (42%) | 13/30 (43%) | 2/6 (33%) |
| Pulmonary<sup>j</sup> | 12/36 (33%) | 11/30 (37%) | 1/6 (17%) |
| Other<sup>k</sup> | 17/36 (47%) | 16/30 (53%) | 1/6 (17%) |
| Any of the above | 23/36 (64%) | 20/30 (67%) | 3/6 (50%) |
| Complications |  |  |  |
| Hospitalization related to complications from product implantation | 9/36 (25%)<sup>c</sup> | 9/30 (30%)<sup>c</sup> | 0 |
| Surgical drainage, debridement, or hardware removal | 4/36 (11%) | 4/30 (13%) | 0 |
| Initial tuberculosis treatment regimen |  |  |  |
| Rifampin, isoniazid, pyrazinamide, ethambutol | 21/36 (58%) | 16/30 (53%) | 5/6 (83%) |
| Rifampin, isoniazid, pyrazinamide | 3/36 (8%) | 2/30 (7%) | 1/6 (17%) |
| Fluoroquinolone-containing regimen | 9/36 (25%) | 9/30 (30%) | 0 |
| Other regimen | 3/36 (8%) | 3/30 (10%) | 0 |
| Treatment outcome |  |  |  |
| Died before completing therapy | 2/36 (6%)<sup>c</sup> | 2/30 (7%)<sup>c</sup> | 0 |
| Completed therapy | 32/36 (89%) | 26/30 (87%) | 6/6 (100%) |
| Stopped therapy early<sup>l</sup> | 2/36 (6%) | 2/30 (7%) | 0 |

<sup>a</sup> Due to rounding, column percentages for initial tuberculosis treatment regimen and outcomes do not sum to 100%.
<sup>b</sup> One spinal recipient who died of culture-confirmed tuberculosis did not undergo an interferon-gamma release assay for *M. tuberculosis* infection.
<sup>c</sup> These counts and percentages exclude one spinal recipient who died from culture-confirmed, disseminated tuberculosis after the end of cohort follow-up.
<sup>d</sup> Imaging abnormalities included surgical site abscesses or fluid collections, osteomyelitis, and discitis; nodular and ground-glass pulmonary opacities and interstitial thickening; basilar meningitis; and brain tuberculomas.
<sup>e</sup> Includes the 11 recipients with microbiologic or imaging evidence of tuberculosis disease.
<sup>f</sup> Cervical, intrathoracic, and inguinal lymph nodes. <sup>g</sup> Surgical site signs and symptoms included erythema, pain, wound drainage, swelling, bleeding, and lymphadenitis. <sup>h</sup> Constitutional signs and symptoms included fever, chills, night sweats, weight loss, fatigue, and loss of appetite. <sup>i</sup> Neurologic signs and symptoms included paresthesia, upper or lower extremity weakness, bowel or bladder dysfunction, dysphagia, headache, confusion, confusion, and altered mental status. <sup>j</sup> Pulmonary signs and symptoms included cough, shortness of breath, and sputum production. <sup>k</sup> Other signs and symptoms included chest pain, nausea, vomiting, diarrhea, anxiety, depression, weakness, dizziness, and photophobia. <sup>l</sup> Two spinal surgery recipients stopped treatment early (after 22 and 94 days of therapy).

Eleven (31%) of the 36 recipients had microbiologic or imaging evidence of tuberculosis disease, all of whom had undergone spinal procedures. Median time from product implantation to first microbiologic or imaging evidence of tuberculosis was 72 days (range, 53–145 days). Anatomic sites of disease included the spine or paraspinal soft tissues (9 recipients [25%]), lungs (4 [11%]), central nervous system (2 [6%]), and lymph nodes (1 [3%]). Spine imaging abnormalities included abscesses or fluid collections (8 recipients) and osteomyelitis or discitis (4 recipients). Pulmonary imaging abnormalities included nodular lesions (2 recipients), ground glass opacities (2 recipients), and interstitial thickening (1 recipient). Brain imaging abnormalities included basilar meningitis and ventriculomegaly (1 recipient) and ring-enhancing nodules consistent with tuberculomas (1 recipient). No resistance to first-line or second-line anti-tuberculosis medications was found on molecular or growth-based testing.

After product implantation, 23 (64%) recipients reported new symptoms consistent with tuberculosis disease. Nine (25%) recipients were hospitalized for complications related to product implantation. Four (11%) underwent additional surgical procedures for infection, including 1 who had the allograft removed 74 days postoperatively.

All 36 recipients started treatment for tuberculosis disease between 25 and 181 days after product implantation (median, 86.5 days) (Supplemental Figure 2). The most common initially prescribed regimen was the standard 4-drug tuberculosis treatment of rifampin, isoniazid, pyrazinamide, and ethambutol (21 [58%] recipients). To optimize bone and central nervous system penetration,^7,8^ some clinicians opted for a regimen that included a fluoroquinolone (9 [25%] recipients).

Two recipients died after 12 and 26 days of treatment (70 and 113 days after spinal surgery, respectively). Both deaths were attributed to tuberculosis. Hematoxylin and eosin staining of a vertebral specimen from one deceased recipient showed bone fragments surrounded by granulomatous inflammation and necrosis. A brain specimen showed features of granulomatous leptomeningitis, and staining with a *Mycobacterium* species immunohistochemical assay demonstrated mycobacterial antigens in areas of granulomatous inflammation.

Two recipients stopped treatment early (after 22 and 94 days of therapy). The remaining 32 recipients completed therapy for tuberculosis disease; their median treatment duration was 288 days (interquartile range, 246–308 days).

### 3.3 Donor investigation

The tissue donor was a man in his 70s born and living in the United States. He was hospitalized after 2 weeks of progressive generalized weakness, dyspnea, cough with yellow sputum production, and confusion. On arrival to the emergency department, the patient’s temperature was 37.1°C, heart rate was 98 beats per minute, blood pressure was 105/58 mmHg, and oxygen saturation was 97% on 4 liters per minute of oxygen. Laboratory evaluation revealed a normal white blood cell count and differential, moderate normocytic anemia, normal platelets, normal renal function, elevated aspartate aminotransferase (approximately 3 times the upper limit of normal), normal alanine aminotransferase, and normal procalcitonin. Initial computed tomography of the chest with intravenous contrast revealed extensive interstitial and ground-glass alveolar opacities bilaterally and a right upper lobe nodule <1 cm in size. He was treated with high-flow oxygen, ceftriaxone, and doxycycline for presumed community-acquired pneumonia. All microbiologic testing was negative, including standard blood, urine, and sputum bacterial cultures, a COVID-19 rapid antigen test, and an influenza A/B rapid PCR test. No mycobacterial testing or testing for *M. tuberculosis* infection was documented in the medical record.

Two weeks after admission, the donor developed worsening encephalopathy. Computed tomography of the head without contrast showed no evidence of acute stroke or another etiology for worsening encephalopathy. Antibiotics were broadened to cefepime and clindamycin, but the patient’s respiratory and mental status continued to worsen. He was placed on comfort-focused care and died. The donor’s underlying cause of death was documented as pneumonia and septic shock.

A standard donor risk assessment interview was conducted with a member of the donor’s household shortly after his death.^9,10^ The interview noted that in recent weeks, the donor had a fever, experienced confusion, and had difficulty walking. The donor also had a chronic cough for years and approximately 8 kg of weight loss in the prior 3 months; the household member attributed the weight loss to the donor’s pneumonia and difficulty swallowing. The household member reported no knowledge of a previous tuberculosis diagnosis, positive testing for *M. tuberculosis* infection, or household exposure to a person with tuberculosis. No tuberculosis risk factors were reported except for age and a remote history of travel of unknown duration to Africa and Central Europe. Standard donor testing for HIV, hepatitis B and C, syphilis, and human T-lymphotropic virus was negative.^11–13^

### 3.4 Tissue processing

Bones (each femur, tibia, fibula, humerus, ulna, radius, and hemipelvis), the tibialis and Achilles tendons, and the fascia latae were recovered from the donor. Only bone products were processed into allograft units for distribution. The bones were transferred to a single tissue establishment, where they were manufactured into a cryopreserved bone matrix product processed to retain live cells.^14^ Following industry practice at that time, standard bioburden testing for bacteria and fungi in samples collected during processing and from units of the final product was negative.^15,16^ In addition, before the product lot was released for distribution, one 2-cc bone allograft unit tested negative for *M. tuberculosis* complex through a laboratory-developed PCR test that the manufacturer voluntarily implemented after the 2021 outbreak.^17,18^

### 3.5 Laboratory testing

After outbreak detection, *M. tuberculosis* was isolated from liquid mycobacterial culture in 2 of 6 unused bone allograft product units that were tested (Figure 1). PCR testing for *M. tuberculosis* remained consistently negative. *M. tuberculosis* isolates from 4 product recipients and 2 unused product units were sequenced and shared a unique whole-genome multilocus sequence type not previously identified in the United States; they differed by 0–1 single nucleotide polymorphism.

### 3.6 Death of product recipient after end of cohort follow-up

A third product recipient died of tuberculosis in 2025, nearly 3 years after receiving the product. During initial evaluation after outbreak detection in 2023, the patient had a positive interferon-gamma release assay but no microbiologic or imaging evidence of tuberculosis disease. She began multidrug treatment for tuberculosis but stopped after 22 doses. In 2025, after beginning immunosuppressive therapy, she developed culture-confirmed, severe, disseminated tuberculosis and died. Tuberculosis was listed as a cause of death. *M. tuberculosis* cultured from this recipient shared the same unique whole-genome multilocus sequence type as the isolates from 4 product recipients and 2 unused product units and differed by 0–2 single nucleotide polymorphisms.

## 4 Discussion

In 2023, a contaminated bone allograft product containing live cells caused the second nationwide tuberculosis outbreak in two years, with substantial morbidity and mortality. Although *M. tuberculosis* concentrations in the allograft appeared low (PCR testing was negative, and only 2 of 6 unused units grew *M. tuberculosis* on culture), transmission was efficient. Among the 36 product recipients, 27 had laboratory evidence of *M. tuberculosis* infection, 11 had clinical evidence of tuberculosis disease, and 2 died of tuberculosis within 12 months of outbreak detection; 1 additional recipient developed fatal tuberculosis nearly 3 years after product receipt. Most surviving recipients underwent 9–12 months of treatment for tuberculosis disease, and four required surgical interventions. Additional details are available in reports published by physicians who cared for these recipients.^3,4^

To prevent disease transmission from contaminated tissue products, FDA policies that prohibit the recovery of tissues from a donor with sepsis should be followed.^11,15^ Adherence to established donor eligibility criteria would have prevented both the 2021 and 2023 outbreaks. Both donors had sepsis of undetermined etiology during terminal hospitalization; the donor in the 2023 outbreak also had pneumonia and chest radiograph findings consistent with (albeit not specific for) tuberculosis disease.^1,20^ The genetic similarity of *M. tuberculosis* isolates from product recipients and unused product units supports the presence of unrecognized tuberculosis disease in the donor at time of death.

Without a processing step that is validated to eliminate contamination with *M. tuberculosis*, the risk of transmission through live-cell tissue products persists. The package insert for the viable bone matrix product in this outbreak described “destructive microbiological testing” and treatment of the “aseptically processed” bone allograft product with povidone iodine, hydrochloric acid, and hydrogen peroxide. However, the manufacturing process was intended to preserve live bone cells, so the bone allograft product itself was not sterile.^14^ Liquid and solid mycobacterial culture testing of tissue products before distribution for transplantation could be considered. However, negative culture results would not eliminate the risk of transmission: in this outbreak, *M. tuberculosis* was isolated in only 2 of the 6 unused bone allograft units that underwent culture testing.

This second multistate tuberculosis outbreak highlights the need for clinicians to closely monitor tissue product recipients and promptly report any adverse events to tissue establishments and public health authorities. Reporting by clinicians of tissue-related adverse health events remains voluntary.^14–16,21^ Fortunately, this outbreak was detected when clinicians in 2 different states independently flagged similarities to the 2021 outbreak and reported their concerns to their respective health departments, with the subsequent public health response averting up to 54 additional procedures with the contaminated product.^1,2^

In September 2024, prompted by this outbreak and others, the federal Advisory Committee on Blood and Tissue Safety and Availability recommended improved communication around informed consent so that patients who receive tissue products understand the potential for communicable disease transmission.^22^ Other recommendations included developing and using electronic biovigilance systems to ensure traceability of tissue products from donors to recipients and back to donors and to enable timely and systematic reporting of adverse events suspected to be due to transmission of communicable diseases by tissue products.^22^

This investigation had two main limitations. The morbidity of this outbreak was likely higher than presented here. Not all product recipients had surgical site imaging or microbiologic testing (i.e., precluding evidence of disease), in part because all recipients were advised to begin tuberculosis treatment even if their diagnostic evaluation was incomplete. The death of one recipient nearly 3 years after receiving the product underscores the potential for delayed morbidity and mortality, especially in the absence of treatment. Another aspect beyond the scope of this report is the extent of additional transmission beyond the 36 recipients. During the 2021 outbreak, 73 healthcare personnel were newly diagnosed with *M. tuberculosis* infection after intraoperative exposure to aerosolized bone allograft product or surgical wound material, postoperative exposure to surgical instruments, or postoperative exposures to recipients and their medical waste (e.g., liquid draining from tuberculous wounds).^23^

In conclusion, prompt detection of this outbreak likely prevented additional morbidity and mortality. Rapid collaboration between CDC, FDA, health departments, and healthcare facilities enabled early tuberculosis treatment initiation for 34 recipients and prevented up to 54 additional procedures with the bone allograft product. Nevertheless, this outbreak demonstrates persistent gaps in tissue transplant safety in the United States. Appropriate donor selection, in conjunction with liquid and solid mycobacterial culture testing of bone products before commercial distribution, could reduce but probably would not eliminate the risk of *M. tuberculosis* transmission. Therefore, it is important that clinicians closely monitor tissue product recipients and promptly report any adverse events to tissue establishments and public health authorities.

## Abbreviations

CDC: (Centers for Disease Control and Prevention)
DNA: (deoxyribonucleic acid)
FDA: (U.S. Food and Drug Administration)
HIV: (human immunodeficiency virus)
*M. tuberculosis*: (*Mycobacterium tuberculosis*)
NAAT: (nucleic acid amplification test)
PCR: (polymerase chain reaction)

## Acknowledgments

Author contributions: KRS, MBH, RJS, PA, SVB, SPA, and JMW conceptualized and designed the investigation. All authors made substantial contributions to the acquisition and interpretation of data. TCT, KAL, JAV, JB, SBS, and LSC performed and interpreted microbiologic and pathologic analyses. KRS, PMW, and NGS performed statistical analyses. KRS, PMW, NGS, and MHB wrote the first draft. All authors critically reviewed subsequent drafts for important intellectual content. All authors reviewed and approved the final manuscript and take full responsibility for all aspects of the work.

The authors thank additional members of the Bone Allograft Tuberculosis 2023 Investigators group for evaluating and treating the affected patients and contacts, locating unused products, and collecting data. Individual names and affiliations are listed in Supplementary Material.

## Data availability

Disclosure of individual patient data from this investigation is prohibited by federal law and CDC policy.

## Use of artificial intelligence (AI) tools

The authors declare that no AI or AI-assisted technologies were used in the writing or editing of this manuscript.

## Supporting information

Additional supporting information may be found online in the Supporting Information section.

