## Supplementary Material for "Second multistate outbreak of tuberculosis caused by a bone allograft product"

**Bone Allograft Tuberculosis 2023 Investigators**

**California**
Mark Beatty — County of San Diego Health and Human Services Agency
Antonio Bejarano — Orange County Public Health Laboratory
Romina Beltrán[a] — County of San Diego Health and Human Services Agency
Zenda Berrada — California Department of Public Health
Yoko Brown — County of San Diego Health and Human Services Agency
Andrea Burger — County of San Diego Health and Human Services Agency
Janet Carrillo-Camacho — County of San Diego Health and Human Services Agency
Debora Cartagena — County of San Diego Health and Human Services Agency
Helene M. Calvet[a] — Orange County Health Care Agency
Yi Ning Cheng — County of San Diego Health and Human Services Agency
Tania Chiem — Orange County Public Health Laboratory
Raymond Chinn — County of San Diego Health and Human Services Agency
Martin Cilnis — California Department of Public Health
Myron Coloma[a] — San Mateo County Health
Coroner Division of Orange County Sheriff's Department
Ashley Cortez[a] — County of San Diego Health and Human Services Agency
Megan Crumpler — Orange County Public Health Laboratory
Camille Dollinger — California Department of Public Health
Erin Epson — California Department of Public Health
Jennifer Flood — California Department of Public Health
Leila Galindo — San Mateo County Health
Lisa L. Goozé[a] — San Mateo County Health
Ankita Kadakia — County of San Diego Health and Human Services Agency
Grace Kang — County of San Diego Health and Human Services Agency
Chris E. Keh[a] — California Department of Public Health
Idamae Kennedy — California Department of Public Health
Janice Kim — California Department of Public Health
Varvara Kozyreva — California Department of Public Health
Aliana Maaghop — County of San Diego Health and Human Services Agency
Nazaneen Mayer — County of San Diego Health and Human Services Agency
Hilary Metcalf — California Department of Public Health
Angie Miner — County of San Diego Health and Human Services Agency
Vivian Nguyen — Orange County Health Care Agency
Jeffrey M. Percak[a] — County of San Diego Health and Human Services Agency
Philip Robinson — Hoag Hospital
Corey Rodriguez — County of San Diego Health and Human Services Agency
Ronabee Rullan-Tangonan — County of San Diego Health and Human Services Agency
Theresa Rymer — County of San Diego Health and Human Services Agency
Yoran (Lana) Sato — California Department of Public Health
Seema Shah — County of San Diego Health and Human Services Agency
Tambi Shaw[a] — California Department of Public Health
Richard Smith — Tri-City Medical Center
Lauralee Spalding — County of San Diego Health and Human Services Agency
Patti Steger — Hoag Orthopedic Institute
Juliet Stoltey — California Department of Public Health

Lawrence Wang — County of San Diego Health and Human Services Agency
Che Waterman — San Mateo County Health
Terry Weber — California Department of Public Health
Wilma Wooten — County of San Diego Health and Human Services Agency
Matthew M. Zahn[a] — Orange County Health Care Agency

**Louisiana**
Oluwafemi Ajibola — Louisiana State University Health Sciences Center
Juzar Ali[a] — Louisiana State University Health Sciences Center
Chris Brown[a] — Louisiana Department of Health
Candace Deshotel — Louisiana Department of Health
Jeanne Gorrondona — East Jefferson General Hospital
Michael Lacassagne[a] — Louisiana Department of Health
Jessica Stapleton[a] — Louisiana Department of Health
Erica Washington — Louisiana Department of Health
Amy W. Wolfe[a] — Louisiana State University Health Sciences Center and Louisiana Department of Health
Crystal Zheng[a] — Tulane University School of Medicine

**Michigan**
Kelly Block — Central Michigan District Health Department
Peter J. Davidson[a] — Michigan Department of Health and Human Services
Robert P. Dickson[a] — Michigan Department of Health and Human Services; University of Michigan
Megan Foster — Munson Healthcare Cadillac Hospital
Daniel R. Kaul[a] — University of Michigan
Annette Marvin — District Health Department #10
Lisa McCormick — District Health Department #10
Jennifer Morse — Mid-Michigan District Health Department and District Health Department #10
Elizabeth Recker — University of Michigan Health – West
Christy A. Scipione — University of Michigan Health
Shona R. Smith[a] — Michigan Department of Health and Human Services
Emily K. Stoneman — University of Michigan Health
Laraine L. Washer — University of Michigan Health

**New Jersey**
Erick Cortes[a] — New Jersey Department of Health
Juliet M. Leonard[a] — New Jersey Department of Health

**New Mexico**
Marcos Burgos[a] — New Mexico Department of Health
RuthAnn Goradia[a] — New Mexico Department of Health
Brenda Montoya Denison[a] — New Mexico Department of Health

**New York**
Christine Chuck — New York City Department of Health and Mental Hygiene
Dawn Cummins — New York City Department of Health and Mental Hygiene
Hannah T. Jordan[a] — New York City Department of Health and Mental Hygiene
Diana M. Nilsen[a] — New York City Department of Health and Mental Hygiene
Brendan Oram — New York City Department of Health and Mental Hygiene

Magdalene Spencer[a] — New York City Department of Health and Mental Hygiene
Jeanne Sullivan Meissner — New York City Department of Health and Mental Hygiene
Lisa Trieu — New York City Department of Health and Mental Hygiene

**Oregon**
Kiley Ariail[a] — Oregon Public Health Division
Heidi Behm[a] — Oregon Public Health Division
Lisa Chambliss — Lane County Health and Human Services
Trevor Hostetler — Washington County Health and Human Services
Patrick Luedtke — Lane County Health and Human Services
Gloria Matthews — Washington County Health and Human Services
Heather Young — Lane County Health and Human Services

**Texas**
Lisa Y. Armitige[a] — University of Texas at Tyler Health Science Center
Amanda Decimo[a] — Texas Department of State Health Services
Nicole Evert — Texas Department of State Health Services
Elizabeth Foy — Texas Department of State Health Services
Annett R. Gonzalez[a] — City of El Paso Department of Public Health
Loretta Hernandez — City of El Paso Department of Public Health
Enid LeBlanc — The Hospitals of Providence
Saroj Rai — Texas Department of State Health Services
Gretchen Rodriguez[a] — Texas Department of State Health Services
Dean E. Smith — El Paso Spine Center

**Virginia**
Leah Breitung — Virginia Department of Health
Kendall Cook — Virginia Department of Health
Sumac Diaz — Virginia Department of Health
Laurie Forlano — Virginia Department of Health
Shania Gupton — Virginia Department of Health
Alexander Samuel — Virginia Department of Health
Vicki Stamps — Virginia Department of Health
Elizabeth Vega — Virginia Department of Health
Melissa Williams — Virginia Department of Health
Laura R. Young[a] — Virginia Department of Health

**U.S. Department of Agriculture (USDA)**
Kimberly A. Lehman[a] — National Veterinary Services Laboratories, Veterinary Services, Animal and Plant Health Inspection Service, U.S. Department of Agriculture
Tyler C. Thacker[a] — National Veterinary Services Laboratories, Veterinary Services, Animal and Plant Health Inspection Service, U.S. Department of Agriculture

**Centers for Disease Control and Prevention (CDC)**
Leeanna Allen — Division of Tuberculosis Elimination (DTE), National Center for HIV, Viral Hepatitis, STD, and TB Prevention (NCHHSTP), CDC
Sandy P. Althomsons[a] — DTE, NCHHSTP, CDC

Pallavi Annambhotla[a] — Division of Healthcare Quality Promotion (DHQP), National Center for Emerging and Zoonotic Infectious Diseases (NCEZID), CDC
Sridhar V. Basavaraju[a] — DHQP, NCEZID, CDC
Elizabeth M. Beshearse — DHQP, NCEZID, CDC
Julu Bhatnagar[a] — Division of Healthcare of High-Consequence Pathogens and Pathology, National Center for Emerging and Zoonotic Infectious Diseases, CDC
Bruce Bradley — DTE, NCHHSTP, CDC
Gail Burns-Grant — DTE, NCHHSTP, CDC
Tracina Cropper — DTE, NCHHSTP, CDC
Lauren S. Cowen[a] — DTE, NCHHSTP, CDC
Justin Davis — DTE, NCHHSTP, CDC
Vanessa Fong — DTE, NCHHSTP, CDC; assigned to California Department of Public Health
Mari Galvis — DTE, NCHHSTP, CDC; assigned to New York City Department of Health and Mental Hygiene
Janet Glowicz — DHQP, NCEZID, CDC
Neela D. Goswami[a] — DTE, NCHHSTP, CDC
Isabel S. Griffin — DHQP, NCEZID, CDC
Marissa K. Grossman — Epidemic Intelligence Service, CDC; assigned to DHQP, NCEZID, CDC
Maryam B. Haddad[a] — DTE, NCHHSTP, CDC
Cam-Van Huynh — Epidemic Intelligence Service, CDC; assigned to DHQP, NCEZID, CDC
David Kuhar — DHQP, NCEZID, CDC
Adam J. Langer — DTE, NCHHSTP, CDC
Philip A. LoBue[a] — DTE, NCHHSTP, CDC
Andrea Lomeli[a] — DTE, NCHHSTP, CDC; assigned to County of San Diego Health and Human Services Agency
Clint McDaniel — DTE, NCHHSTP, CDC
Emily McDonald — DHQP, NCEZID, CDC
Scott A. Nabity[a] — DTE, NCHHSTP, CDC; assigned to California Department of Public Health
Michele Neuburger — Division of Oral Health, National Center for Chronic Disease Prevention and Health Promotion, CDC
Kiran Perkins — DHQP, NCEZID, CDC
Joseph Perz — DHQP, NCEZID, CDC
Shameer Poonja — DTE, NCHHSTP, CDC
Shanica Railey — DTE, NCHHSTP, CDC
Kala Marks Raz — DTE, NCHHSTP, CDC
Sarah Reagan-Steiner[a] — Division of Healthcare of High-Consequence Pathogens and Pathology, National Center for Emerging and Zoonotic Infectious Diseases, CDC
Caitlin Reed — DTE, NCHHSTP, CDC
Paul Regan — DTE, NCHHSTP, CDC
Frank Romano — DTE, NCHHSTP, CDC; assigned to New Jersey Department of Health
Audilis Sanchez[a] — DTE, NCHHSTP, CDC; assigned to Texas Department of State Health Services
Cassandra Schember — Epidemic Intelligence Service, CDC; assigned to California Department of Public Health
Kimberly R. Schildknecht[a] — Epidemic Intelligence Service, CDC; assigned to DTE, NCHHSTP, CDC
Noah G. Schwartz[a] — DTE, NCHHSTP, CDC
Julie Self — DTE, NCHHSTP, CDC
Angela M. Starks[a] — DTE, NCHHSTP, CDC
Rebekah J. Stewart[a] — DTE, NCHHSTP, CDC

Sarah Talarico — DTE, NCHHSTP, CDC
Julian A. Villalba[a] — Division of Healthcare of High-Consequence Pathogens and Pathology, National Center for Emerging and Zoonotic Infectious Diseases, CDC
Paula M. Williams[a] — Epidemic Intelligence Service, CDC; assigned to DTE, NCHHSTP, CDC
Jonathan M. Wortham[a] — DTE, NCHHSTP, CDC

[a]Member of writing group.

### Supplementary Methods

Real-time Polymerase Chain Reaction (rt-PCR) and Culture

Approximately 100 mg of the bone allograft material was transferred to a screw-top tube containing 400 µL 1x TE with 100 µL 1.0 mm and 200 µL 0.1 mm glass beads, along with an extraction control. The sample was heat-inactivated and then homogenized using a BioSpec Beadbeater as recommended by the manufacturer. Deoxyribonucleic acid (DNA) was isolated, and the presence of *M. tuberculosis* complex DNA was detected using a real-time polymerase chain reaction (PCR) assay that amplifies the IS*1081* insertion element. The remaining material was homogenized in 7 mL sterile phosphate-buffered saline (PBS) using a gentleMacs Tissue Homogenizer. The soluble portions of the samples were centrifuged to pellet any potential mycobacteria in the sample. The supernatant was decanted, and the pellet resuspended in 3 mL sterile PBS. 400 µL was used to inoculate BD BACTEC Mycobacterial Growth Indicator Tube (MGIT) media in duplicate. 1.5 mL of 800 µg/mL erythromycin was added to the MGIT PANTA dry mix supplied in the BACTEC MGIT Supplement Kit. Then, 15 mL of the reconstituting fluid was added to the PANTA/erythromycin mix. 0.8 mL of the reconstituted PANTA/erythromycin mix was added to each MGIT tube. Using a swab, commercially available solid media (Stonebrinks, Lowenstein-Jensen, and Mycobactosel-Lowenstein-Jensen) and in-house prepared solid media (7H11 with pyruvate and 7H11 with glycerol) were inoculated.

Whole-Genome Sequencing and Phylogenetic Analysis

Nextera XT sequencing libraries were sequenced (2 x 150 bp reads) on the NextSeq 500 platform using NextSeq 500/550 Mid Output Kit v2.5 300 cycle chemistry. Samples with a sequence read set Q30 frequency <85% were re-sequenced. Reference-guided assemblies were created using BioNumerics 7.6 Reference Mapper v 1.2.3 using *M. tuberculosis* strain H37Rv (NC00962.3) as the reference with the following settings for base calling: minimum total coverage=3, minimum forward coverage=1, minimum reverse coverage=1, single base threshold=0.75, double base threshold=0.85, triple base threshold=0.95, gap threshold=0.5. A bowtie-based algorithm was used.[1] Samples with an average genome coverage <25 were re-sequenced. Reference-guided assembly samples were compared using BioNumerics 7.6 single nucleotide polymorphism (SNP) analysis filters. For a SNP to be retained in the comparison, the base in all samples must have had a total coverage of 5 reads, must not have contained ambiguous bases, must not have contained unreliable bases, must not have contained gaps, and must not have been within 12 base-pairs of another SNP. SNPs that were non-informative (identical in all samples) were also excluded.

**Supplementary Figure 1. Timeline of Second Multistate Outbreak of Tuberculosis Caused by Surgical Implantation of a Bone Allograft Product, February to July 2023**

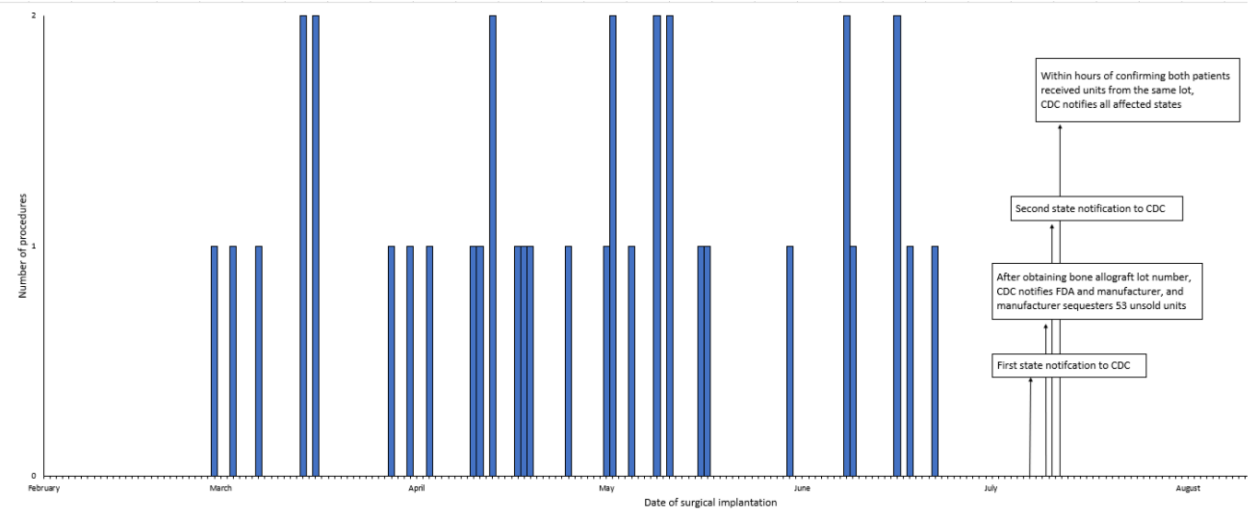

Bone was procured from a deceased donor and processed into a cryopreserved bone matrix product that retained live cells. Fifty units were distributed to 13 healthcare facilities (8 hospitals and 5 dental facilities) in 7 U.S. states during February 27–June 20, 2023. During February 28–June 22, 2023, 49 units were implanted into 36 recipients who resided in 9 states.

On July 7, 2023, a state health department notified the Centers for Disease Control and Prevention (CDC) about a person diagnosed with meningeal tuberculosis 5 weeks after a bone allograft product was used in spinal fusion surgery. During July 8–9, CDC began collecting additional information including the manufacturer name and lot number of the bone allograft product. On July 10, CDC notified the U.S. Food and Drug Administration (FDA) and the product manufacturer (i.e., tissue establishment) about the tuberculosis case. Each lot number corresponds to one unique donor. The product manufacturer immediately sequestered (i.e., quarantined and removed from distribution) all units from the product lot that had not already been distributed for use. On July 11, the product manufacturer also provided CDC and FDA with sales records of hospitals and dental facilities that had already received units from this product lot. On the same day, another state health department (unaware of the first notification) notified CDC about a patient with disseminated tuberculosis after a bone allograft product was used in back surgery. On July 12, the second state health department reported that the second patient's surgical procedure used bone allograft product with the same lot number, confirming that the 2 patients in both states had received tissue from the same donor. Within hours, CDC used the sales records provided by the product manufacturer after the first notification to notify each of the affected 7 state health departments with a line list of each unit distributed to a hospital or dental facility within that state. Concurrently, CDC recommended that all recipients be evaluated and treated with empiric multidrug treatment for tuberculosis disease immediately, regardless of signs and symptoms.

Alt text: Timeline depicting the rapid efforts by public health authorities to evaluate product recipients and sequester unused product after two tuberculosis cases were independently reported among product recipients in July 2023.

**Supplementary Figure 2. Days Between Product Implantation and Start of Treatment for Tuberculosis Disease, by Patient Outcome**

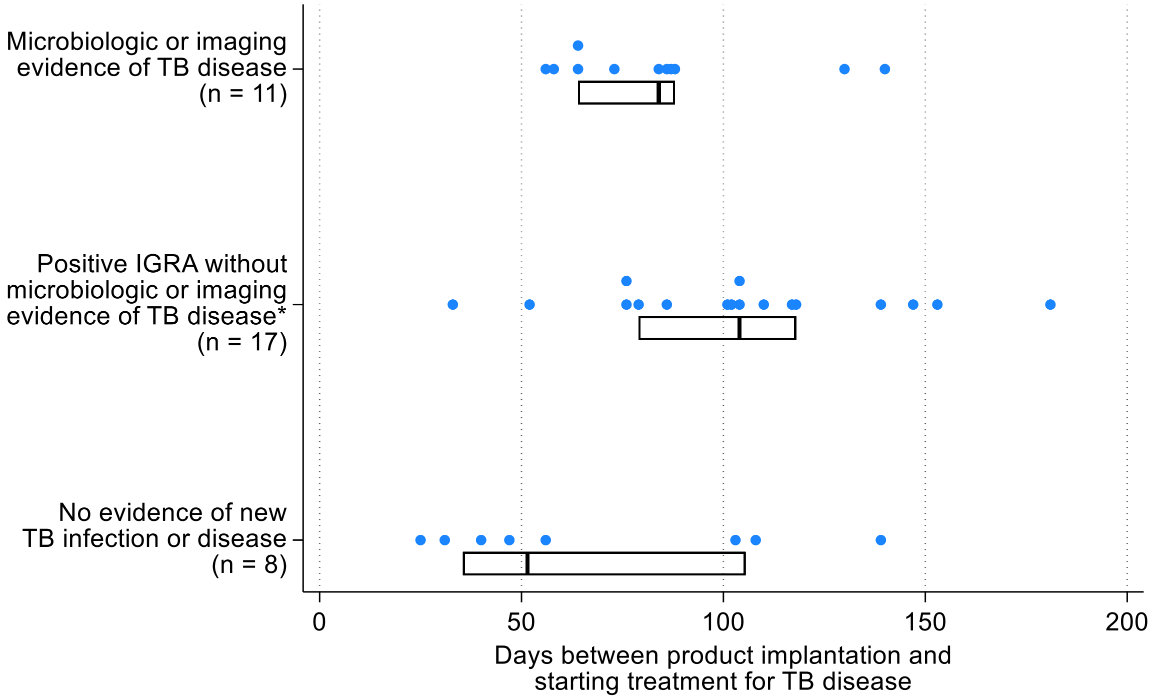

Abbreviations: IGRA, interferon-gamma release assay; TB, tuberculosis.

Each dot represents one product recipient. Boxes represent the $25^{th}$, $50^{th}$, and $75^{th}$ percentile values. Microbiologic evidence of tuberculosis disease was defined as detection of *Mycobacterium tuberculosis* complex in a clinical specimen by mycobacterial culture or nucleic acid amplification testing, including polymerase chain reaction.

* Some product recipients in the second category reported clinical signs and symptoms consistent with tuberculosis disease (such as cough, night sweats, weight loss) that resolved after they began treatment.

Alt text: Graph comparing the number of days between product implantation and start of tuberculosis treatment for recipients with three outcomes: microbiologic or imaging evidence of tuberculosis disease, a positive interferon-gamma release assay without microbiologic or imaging evidence of disease, and no evidence of tuberculosis infection or disease.
